# The impact of vaccination strategies for COVID-19 in the context of emerging variants and increasing social mixing in Bogotá, Colombia: a mathematical modelling study

**DOI:** 10.1101/2021.08.06.21261734

**Authors:** Guido España, Zulma M. Cucunubá, Juliana Cuervo-Rojas, Hernando Díaz, Manuel González-Mayorga, Juan David Ramírez

**Affiliations:** Department of Biological Sciences and Eck Institute for Global Health, University of Notre Dame, USA; Departamento de Epidemiología Clínica y Bioestadística, Facultad de Medicina, Pontificia Universidad Javeriana, Bogotá, Colombia; MRC Centre for Global Infectious Disease Analysis, J-IDA, Imperial College London, London, UK; Departamento de Ingeniería Eléctrica y Electrónica, Universidad Nacional de Colombia, Bogotá, Colombia; Sub-secretaría de Salud Pública. Secretaría Distrital de Salud de Bogotá, Bogotá, Colombia; Centro de Investigaciones en Microbiología y Biotecnología-UR (CIMBIUR), Facultad de Ciencias Naturales, Universidad del Rosario, Bogotá, Colombia

## Abstract

**Background:** In Bogotá by August 1st, more than 27,000 COVID-19 deaths have been reported, while complete and partial vaccination coverage reached 30% and 37%, respectively. Although reported cases are decreasing, the potential impact of new variants is uncertain.

**Methods:** We used an agent-based model of COVID-19 calibrated to local data. Variants and vaccination strategies were included. We estimated the impact of vaccination and modelled scenarios of early and delayed introduction of the delta variant, along with changes in mobility, social contact, and vaccine uptake over the next months.

**Findings:** By mid-July, vaccination may have prevented 17,800 (95% CrI: 16,000 - 19,000) deaths in Bogotá. We found that delta could lead to a fourth wave of magnitude and timing dependent on social mixing, vaccination strategy, and delta dominance. In scenarios of early dominance of delta by mid-July, age prioritization and maintaining the interval between doses were important factors to avert deaths. However, if delta dominance occurred after mid-September, age prioritization would be less relevant, and the magnitude of a four wave would be smaller. In all scenarios, higher social mixing increased the magnitude of the fourth wave. Increasing vaccination rates from 50,000/day to 100,000/day reduced the impact of a fourth wave due to delta.

**Interpretation:** The magnitude and timing of a potential fourth wave in Bogotá caused by delta would depend on social mixing and the timing of dominance. Rapidly increasing vaccination coverage with non-delayed second doses could reduce the burden of a new wave.

**Funding:** NSF RAPID DEB 2027718. HERMES 50419. Medical Research Council. MR/R024855/1

**Research in Context:** *Evidence before this study:* The impact of vaccination strategies in the context of emerging SARS-CoV-2 variants and increasing social mixing in Colombia had not been previously evaluated through mathematical modelling. We searched PubMed for modelling studies using the terms “COVID-19 vaccine AND model AND variant AND Colombia” or “SARS-CoV-2 AND vaccine AND model AND variant AND Colombia” (From 2021/1/1 to 2021/07/31). We did not find studies addressing this question. However, we found a model describing the evolution of the epidemic in the country during the first year, and research on the emergence of alpha, gamma, and B.1.621 variants in Colombia. We extended a previous version of our SARS-CoV-2 agent-based model for Bogotá to include the potential effect of vaccination and variants. This model simulates transmission of SARS-CoV-2 based on daily activity patterns of a synthetic population, representing demographic and geographic characteristics of the total population of the city.

*Added value of this study:* First, our study provides a preliminary estimate of the impact of the vaccination program in Bogotá in terms of the number of deaths prevented. The second major finding is the indication that due to the introduction of the delta variant in the city, and based on the current knowledge of its biology, there is a risk of a fourth epidemic wave, whose time of occurrence and magnitude would depend mainly on three factors: when delta becomes dominant, the intensity of social contact, and vaccination roll-out strategy and coverage.

*Implications of all the available evidence:* We estimate that by mid-July, vaccination may have already prevented 17,800 (95% CrI: 16,000 - 19,000) deaths in Bogotá. The delta variant could become dominant and lead to a fourth wave later in the year, but its timing will depend on the date of introduction, social mixing patterns, and vaccination strategy. In all scenarios, higher social mixing is associated with a fourth wave of considerable magnitude. If an early delta introduction occurred (dominance by mid-July), a new wave may occur in August/September and in such case, age prioritization of vaccination and maintaining the 21-day interval between doses of the Pfizer-BioNTech BNT162b2 are more important. However, if introduction occurred one or two months later (with dominance by mid-August/September) a fourth wave would be of smaller magnitude, the age-prioritization is less relevant, but maintaining the dose scheme without postponement is more important. In all scenarios we found that increasing the vaccination rate from the current average of 50,000/day to 100,000/day reduces the impact of a potential fourth wave due to the delta variant. Our study indicates that given the possibility of a fourth wave in the city, it is necessary to continue maintaining adherence to non-pharmacological interventions, such as the use of face masks and physical distancing, to be cautious with the intensification of social activities, and that it is essential to increase the current pace of vaccinations to rapidly reach high vaccination coverage in the population of the city.

## Introduction

In Colombia, by the beginning of August 2021, 4.8 million COVID-19 cases and 122,000 deaths due to COVID-19 had been reported [1]. Bogotá contributes to close to 30% of the cases in the country, with 1.4 million reported by the first week of August [2]. The third wave of the epidemic in the country surpassed the magnitude of the previous two, and overwhelmed the capacity of the health system.

Vaccination against COVID-19 initiated in the country on February 17, 2021 with various vaccines introduced progressively (CoronaVac/Sinovac, Pfizer-BioNTech BNT162b2, Oxford/AstraZeneca (AZD1222), Janssen (J&J) Ad26.COV2.S, and more recently Moderna (mRNA-1273). The Ministry of Health defined a prioritization strategy that started with health care workers, and then in the general population beginning with the older age groups and those with comorbidities [3]. By the beginning of August, 32% of the total population of Colombia had received at least one dose, and about 25% had a complete scheme [3,4]. In Bogotá, 5.3 million doses had been administered, with 38% of the population having received at least one dose, and close to 30% a completed vaccination scheme [5]. Under the age-prioritization strategy, the country has started to vaccinate those above 25 years[5].

Various SARS-CoV-2 variants have been reported in Colombia. The alpha variant (B.1.1.7) was first reported on April 16, 2021, and the gamma variant (P.1) on January 29, 2021. In addition, several regions in Colombia have reported since the beginning of January the B.1.621 variant, which then became dominant during the third wave of COVID-19 [6]. The delta variant (B.1.617.2) was reported on July 24, 2021. This variant of concern (VOC) has overtaken as dominant in various countries in Latin America such as Brazil, Mexico, and Costa Rica [7]. Delta has been described in the United Kingdom as 1.5 times more transmissible than alpha, with increased severity, and reducing the effectiveness of vaccines [8,9]. Given the characteristics of delta, it is important to quantify its potential impact if it becomes the dominant variant in Bogotá.

We used a detailed agent-based model of the epidemic in Bogotá [10,11] to explore the impact of multiple circulating variants, including delta. We also studied the impact of changing social mixing patterns in the city and of different vaccination strategies.

## Results

### Model calibration until third wave

The model was calibrated to daily reported deaths and seroprevalence data in Bogotá. Using preliminary data on variants dominance in the city, observed mobility, and vaccination coverage, we reproduced the daily trends of deaths, as well as the effective reproduction number and daily demand of intensive care unit (ICU) beds in the city (Fig 1, Fig S4). Although the third wave has been the largest in the city, and the model shows an attack rate over 80% in the city by July, our simulations show that the vaccination program already had a large impact on the peak and total number of deaths. Up to mid-July 2021, we estimate a total of 17,800 (95% CrI: 16,000 - 19,000) deaths averted by vaccination (Fig 1).

**Fig 1.**
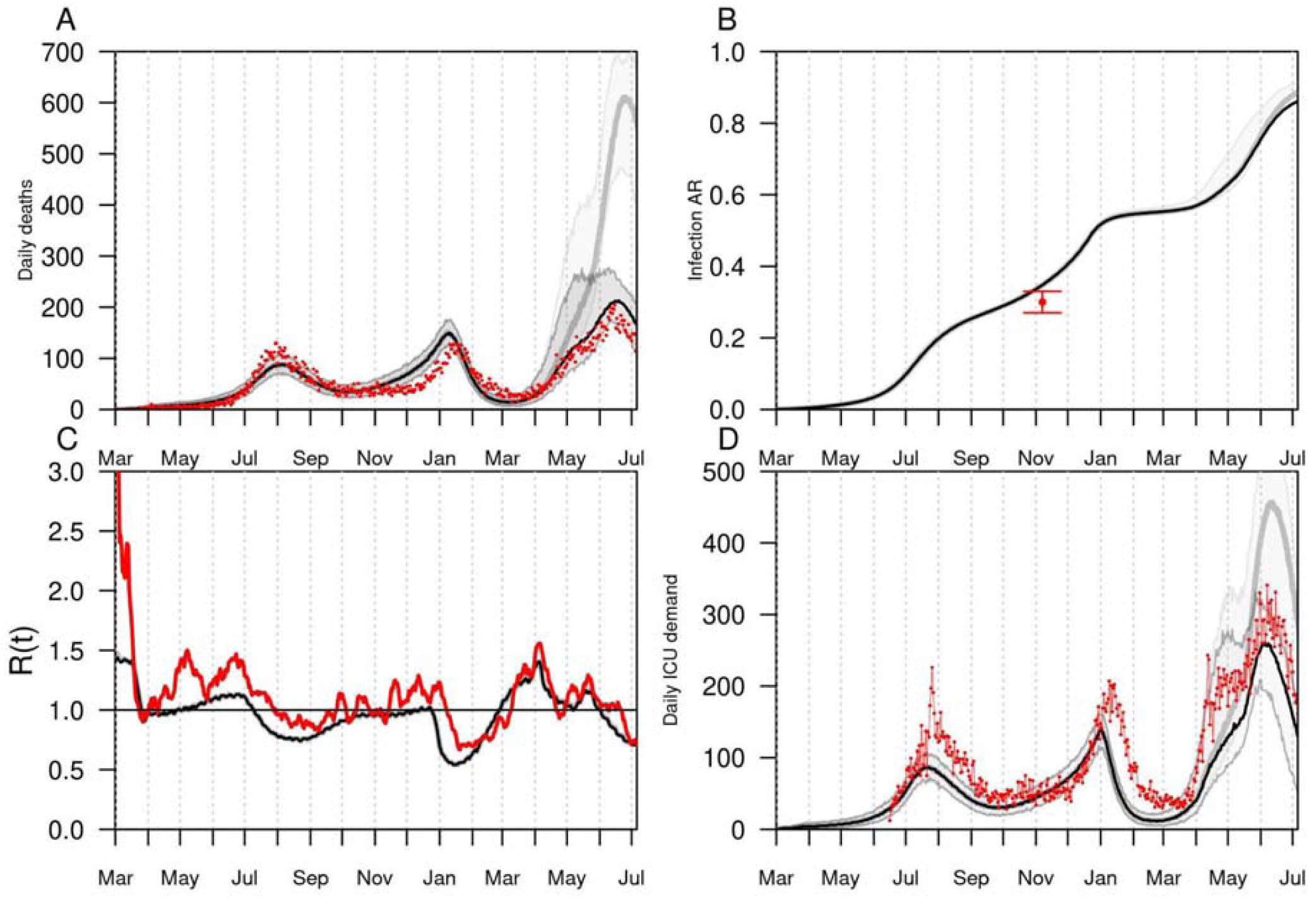
Model calibration and validation until third SARS-CoV-2 wave in Bogotá, Colombia. Panel A shows the daily number of deaths in the city, panel B shows the estimated infection attack rate, panel C shows the effective reproductive number, and panel D shows the daily demand for intensive care unit beds (ICU) due to COVID-19. In all panels, black lines show the calibrated model and red color the data. The gray lines show the dynamics of an alternative scenario without vaccines.

The model results suggest that the third wave of infections was caused mainly by the B.1.621 followed by the gamma variant (Fig S3). We estimated that the B.1.621 variant is likely to be 1.2 (1.2 -1.98) times more transmissible and able to evade immunity from previous infections at 37% (19% - 48%), compared to the original lineage. We estimated the potential early introduction of the alpha variant in Bogotá to be around November 2020, followed by the introduction of gamma early in January, 2021 (Fig S2), whereas the appearance of B.1.621 may have occurred later in the month [6]. Based on this framework, our baseline scenario assumed that importations from the delta variant may have started in Bogotá since May 2021 (Fig S2), but in such a case, delta would not become dominant before August (Fig S3). Calibrated parameters are listed in Table S3.

### Model future projections

The baseline model assumes that the delta variant has already been introduced in the city but it is not yet dominant. Projections show a fourth wave of COVID-19 in the city due to the delta variant, however, its magnitude would depend on the date of introduction of delta, the social mixing patterns, and the vaccination coverage achieved at the time of dominance (Fig 2).

**Fig 2.**
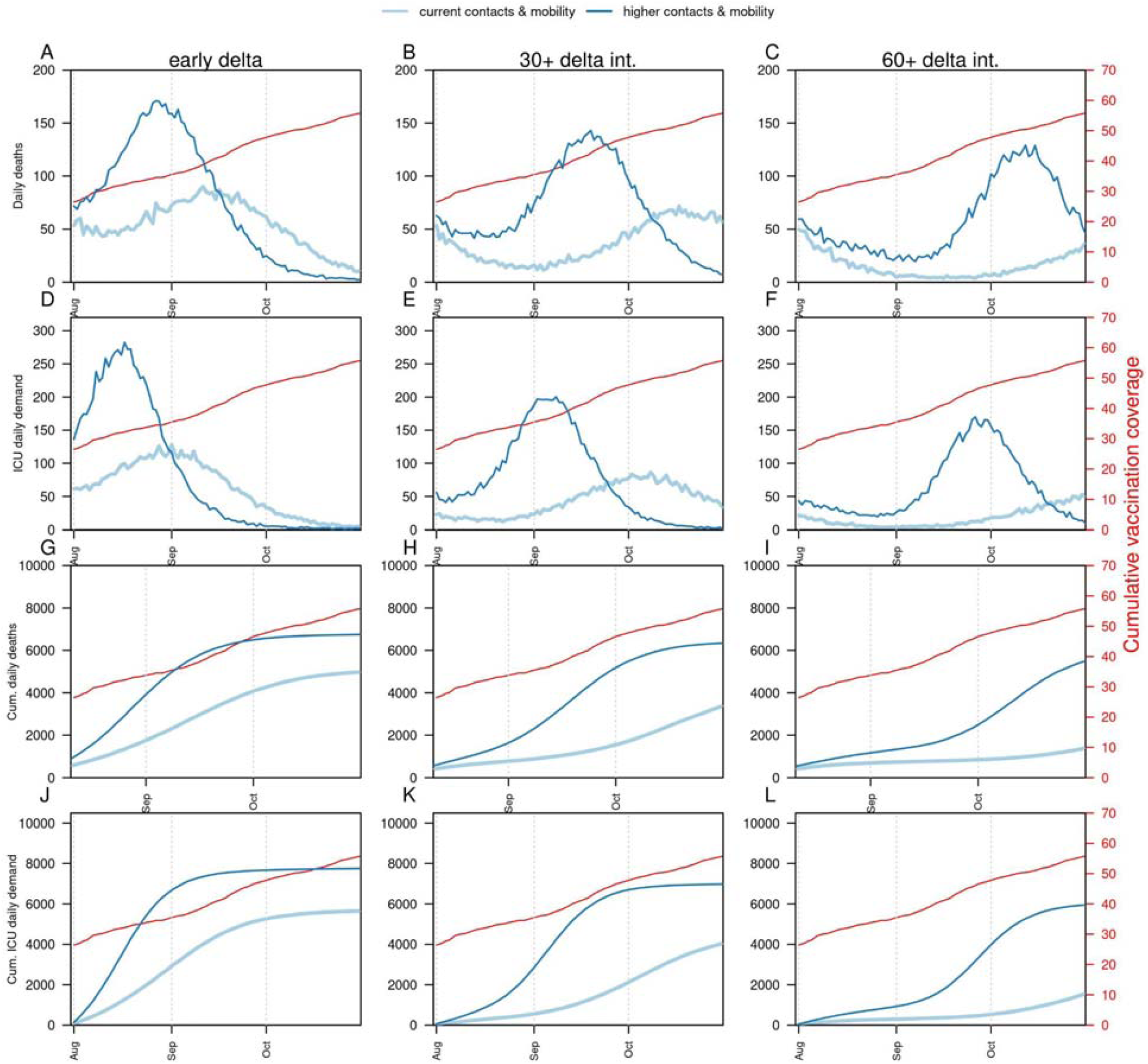
Projections of potential impact on deaths and ICU demand of delta variant on a fourth SARS-CoV wave in Bogotá, according to timing of delta introduction and social mixing patterns. Columns show the timing of delta introduction, defined as early (calibrated), 30+ delta (delayed 30 days), and 60+ delta (delayed 60 days). Panels A-C show the daily number of deaths, panels D-F show the daily demand for ICU beds, panels G-I show the cumulative number of deaths from August to November, and panels J-L show the cumulative number of ICU beds demanded in the city from August to November. Dark blue lines show a scenario of high mobility and high contacts, and light blue lines indicate the current estimated levels of mobility and contacts in the city. The red lines (right axis) represent the coverage of full vaccination in proportion to the total population.

Our model results show that the magnitude of a fourth wave of COVID-19 in Bogotá would depend on the intensity of social mixing over the coming months. The current level of social mixing assumes a moderate-high level of mobility with a moderate level of contacts, while the scenario of higher social mixing assumes an increase in the number of contacts per person, in the community overall, in addition to high levels of mobility. In all scenarios, higher social mixing increased the projected number of deaths and ICU demand in the context of a dominant delta variant (Fig 2). With high social mixing, an early introduction of delta would result in a peak of 189 (95% CrI: 160-245) daily deaths, whereas a late introduction would cause a peak of 130 (95% CrI: 116 - 180). In both scenarios of the introduction of delta, reduced social mixing resulted in a smaller peak. With moderate social mixing, an early introduction of delta would result in a peak of 104 (95% CrI: 77-150) daily deaths, compared to 59 (95% CrI: 47-136) daily deaths in the scenario of late introduction of delta.

Another determinant of the potential magnitude of the fourth wave is the vaccination coverage achieved by the time delta is dominant. In scenarios of an early introduction of delta (dominance in August), our simulations showed that a new wave may occur in August/September, and in such case, changes in the vaccination strategies would have a small effect in the dynamics of the fourth wave. Nonetheless, increasing the vaccination rate to 100,000 vaccines/day could slightly reduce the impact on deaths and ICU demand if age continues to be prioritized and the interval between doses of the Pfizer-BioNTech BNT162b2 is not extended to 84 days. Extending the dose interval to 84 days resulted in an increased number of infections and deaths in this scenario, even with higher vaccination rates (Fig 3 & S5, left column). In the scenario in which delta introduction occurs two months later (dominance in October), the age-prioritization for adults is less relevant but maintaining the dose scheme without postponement is more important, given that those with only one vaccine dose have lower protection against infection, particularly for delta (Table S1), which resulted in an increased number of infections and deaths at the population level (Figs S6, S7). In the scenario of dominance of delta in October, we found that increasing the vaccination rate to 100,000/day (the maximum possible according to health authorities) could have a larger impact in reducing the burden of a fourth wave due to the delta variant. These findings are more pronounced if assuming that efficacy against infection is 100% of the efficacy against symptoms (Figs S8 & S9).

**Fig 3.**
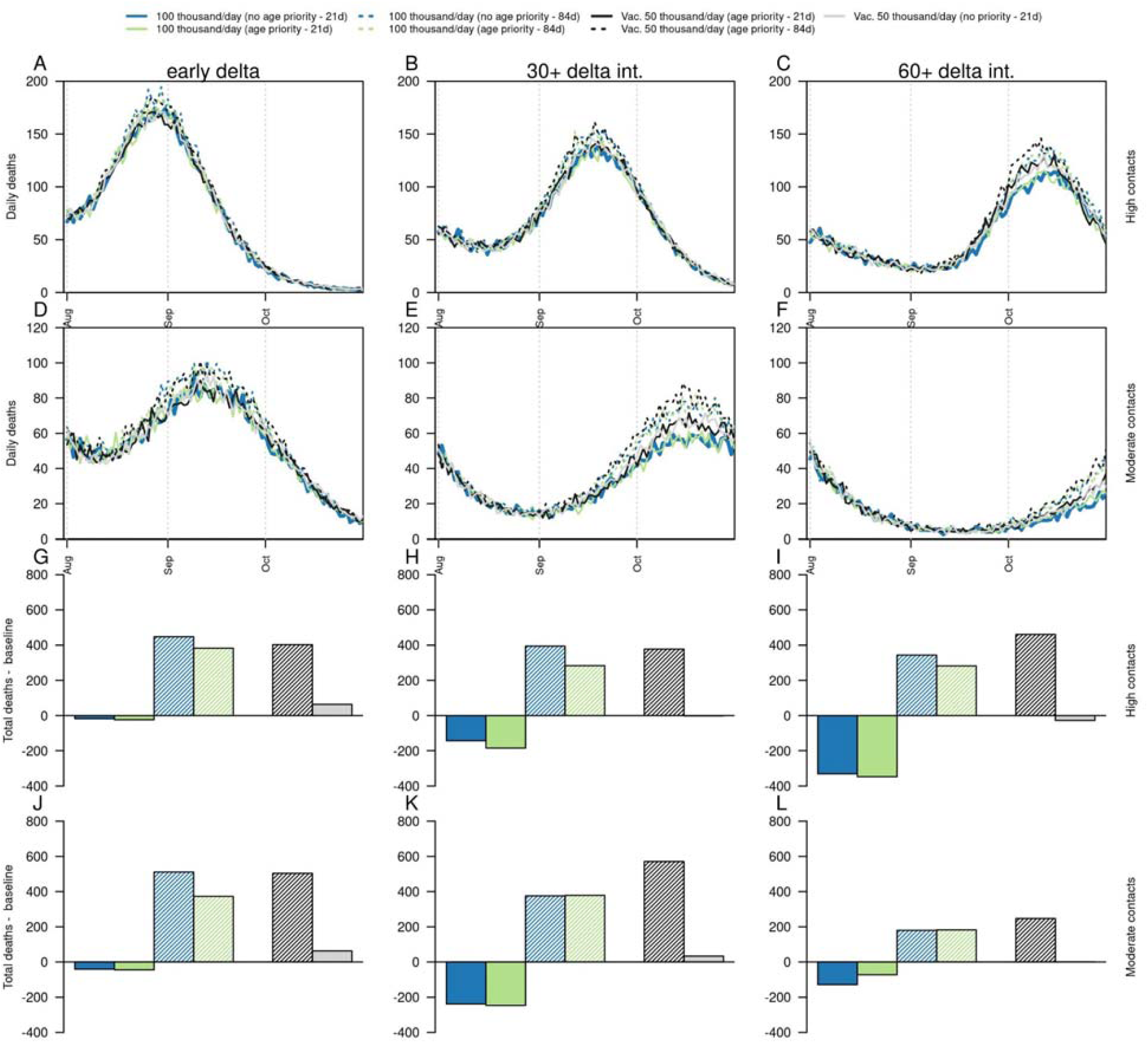
Projections of the potential impact on deaths of delta variant on a fourth SARS-CoV-2 wave in Bogotá, according to timing of delta introduction, level of social mixing, and vaccination strategies. Columns show the timing of delta introduction, defined as early (calibrated), 30+ delta (delayed 30 days), and 60+ delta (delayed 60 days). Panels A-C show the daily number of deaths under a scenario of high social mixing, panels D-F show the daily number of deaths under a scenario of moderate social mixing, panels G-I show the difference in the cumulative number of deaths between alternative vaccination strategies and the baseline scenario (50 thousand vaccines/day with age prioritization and non-postponed second dose of the Pfizer vaccine) with high social mixing, and panels J-L show the difference in the cumulative number of deaths between alternative vaccination strategies and the baseline scenario (50 thousand vaccines/day with age prioritization and non-postponed second dose) with moderate social mixing. Black line shows the baseline scenario of mobility and current vaccination strategy. Dashed lines show the impact of increasing the interval between doses of the Pfizer vaccine to 84 days. Blue colors show the impact of increased vaccination rates (100,000/day) without age priority. Green colors show the impact of increased vaccination rates (100,000/day) with age priority. Gray colors show the impact of baseline vaccination rates without age priority.

## Discussion

We simulated the impact of the introduction of the delta variant using an agent-based model that includes a detailed representation of the population of Bogotá by age, geographic location, and main social activities and mobility patterns (schools, universities, workplaces, long-term care facilities, households, and neighborhoods). This model has been previously validated to COVID-19 dynamics in various places [10,11]. We found that the increased number of cases and deaths during the third wave of COVID-19 in the city could be explained by a combination of higher mobility and social contacts, along with the presence of variants of concern or interest in particular gamma may explain the first part of the third wave, whereas B.1.621 the second part. B.1.621 may have overcome the gamma variant as dominant despite being potentially introduced at similar times. Interestingly, we found that the alpha variant was not a driver of the third wave. Our model suggests that despite an estimated high infection attack rate in the city, that there is still a risk of an additional fourth wave produced by the delta variant, whose amplitude would depend on the timing of dominance of delta, the vaccination coverage and strategy, and the level of social mixing over the next months.

We evaluated the potential impact of postponing the second dose of the Pfizer-BioNTech BNT162b2 vaccine on the potential future dynamics of COVID-19 in the city given delta dominance. We found that in our context this may not contribute to reducing the impact of a fourth wave, in contrast to previous modeling analyses that have shown benefits at the population level [12,13]. However, those studies have been evaluated for non-delta variants. Also, the benefits of second dose postponement is still under debate [14,15]. Some studies have suggested that postponing the second dose may be related with stronger and more durable immune protection [16,17]. However, even if this holds, postponement of the second dose would result in less protection against infection and disease at the individual level for those with an incomplete vaccination scheme under the imminence of the presence of delta variant, which may result in a higher number of infections, and consequently deaths, at the population level. Indeed, recent studies indicate that the effectiveness of the first dose of the Pfizer-BioNTech BNT162b2 and the Oxford/AstraZeneca (AZD1222) vaccines considerably decreases for the delta variant (36% and 30%, respectively) [8].

We also evaluated the impact of different vaccination roll-out strategies. We found that even if vaccination rates are doubled, under a scenario of early introduction of delta, there might not be enough time to mitigate the impact of a fourth wave. However, if the introduction of delta is delayed or social mixing remains moderate, increasing the vaccination rates may result in a milder fourth wave. Our results suggest that in the context of a delayed presence of delta, a preferred strategy for the city is maintaining moderate levels of social mixing combined with a rapid increase in vaccination rates (even without age prioritization) and administering the second dose of the Pfizer-BioNTech BNT162b2 vaccine without postponement.

Based on data available on vaccination coverage and vaccine hesitancy for adults being estimated in population-based surveys at 12,8% [18], we assumed that about 10% of the target population remains not vaccinated, despite already being eligible. This means that for each target group defined by the vaccination campaign roll-out plan there is a remaining high-risk population that has not been vaccinated (due to hesitancy or access barriers). Although we did not model strategies to reach these populations, vaccinating these groups will have the highest impact in reducing severe disease outcomes in a future wave of COVID-19.

### Limitations and strengths

Our model relies on data to adjust current dynamics and to project hypothetical scenarios of future transmission. Currently, there is scarce data corresponding to dominance of SARS-CoV-2 variants in the city, which may affect our estimates of dominance of variants. Also, our projections rely on our ability to project future social mixing patterns, which has proved challenging. For these reasons, we have considered scenarios of moderate and high social mixing levels over the coming months. Our model also relies on available data on vaccination efficacy or effectiveness against multiple variants. However, for some of them, there is still considerable uncertainty. These include, for instance, the effectiveness of CoronaVac/Sinovac vaccine against the delta variant, and of all the vaccines against the B.1.621 variant. Moreover, our model has not considered data on waning of immunity over time, but we consider parameters for loss of immune protection against infection after natural infection as a fixed parameter, according to literature available at the time. Also, our model assumes vaccine efficacy as a parameter but not many vaccine efficacy estimates against different variants are available and for that reason in some cases we have used vaccine effectiveness as a proxy for vaccine efficacy. We have also used estimates of vaccine efficacy against variants developed by other authors (Table S1). Importantly, we have considered differential progression parameters for the original wild type virus and alpha and delta variants as reported in the literature. However, there is no information on those parameters for other variants. Another aspect not yet considered in our model is whether vaccination after previous infection may provide higher protection, or whether the efficacy of vaccines varies considerably according to characteristics of individuals, such as age or comorbidities [19]. This is particularly important in our context as we project a high attack rate before mass vaccination started. However, at the time of writing this manuscript, there is no information to parameterize these variations in efficacy within the model. Further work may also include estimates of the potential impact of boosters, particularly in high-risk populations.

## Data Availability

Data used in this project is publicly available. The code of this project can be accessed on https://github.com/confunguido/FRED

## Acknowledgements

We thank the Secretaría de Educación Distrital de Bogotá and Secretaría de Cultura for providing information and valuable insights into schools and universities distribution, size and demographics. We thank health professionals at Secretaría Distrital de Salud de Bogotá and SaluData for providing daily updated information on COVID-19 cases, deaths, ICU occupancy and demand. We thank the University of Notre Dame Center for Research Computing for computing resources. GE received funding from an NSF RAPID grant (DEB 2027718). HD received partial funding from the National University of Colombia (Universidad Nacional de Colombia (HERMES 50419)). ZMC receives funding from the Medical Research Council (MR/R024855/1). ZMC and GE are *ad honorem* members of the Scientific Advisory Group on epidemiological modeling at Secretaría de Salud of Bogotá. MGM holds a decision-making position at Secretaría de Salud of Bogotá. We thank Charlie Whittaker from MRC-GICA at Imperial College London for his valuable input into variant characteristics and modeling. The funding sources had no involvement in the collection, analysis, interpretation of data, the writing of the report, or in the decision to submit the paper for publication.

## Methods

### Data

Time-varying trends of cases and deaths were obtained from public data sources. We used the daily number of deaths reported in Bogotá as of mid-July 2021 [20], population based seroprevalence data as of Nov, 2020 (based on anti-SARS CoV-2 IgG) [21], and available prevalence data on variants over the third wave in Bogota based on 120 samples from the epidemiological weeks 12 to 22 [22] to calibrate the model parameters. To represent changes in mobility, we used Google Mobility Reports [23] and Grandata project [24], both freely available online. Data on vaccination were obtained from datasets available online from the Ministry of Health [25](Fig S1). We obtained daily ICU bed demand from the Secretary of Health of Bogotá. Other data sources to create the synthetic population were obtained from official population projections by age for 2021[26], IPUMS-International [27] and the Secretary of Education of Bogotá.

### Model

We extended a previous version of our SARS-CoV-2 model for Bogotá [10] to include the potential effect of vaccination and variants. Our agent-based model simulates transmission of SARS-CoV-2 based on daily activity patterns of a synthetic population (Fig. 4), representing demographic and geographic characteristics of the total population of Bogotá. Social mixing was considered in the model assuming two ways to increase contacts: mobility and average number of contacts. Mobility in the model represents the proportion of people who leave their household to participate in other activities, such as school or work. Community contacts are included in the model as an average number of contacts that an individual has in the community, given that the individual leaves the household. For instance, both mobility and community contacts may increase in holidays.

**Fig 4.**
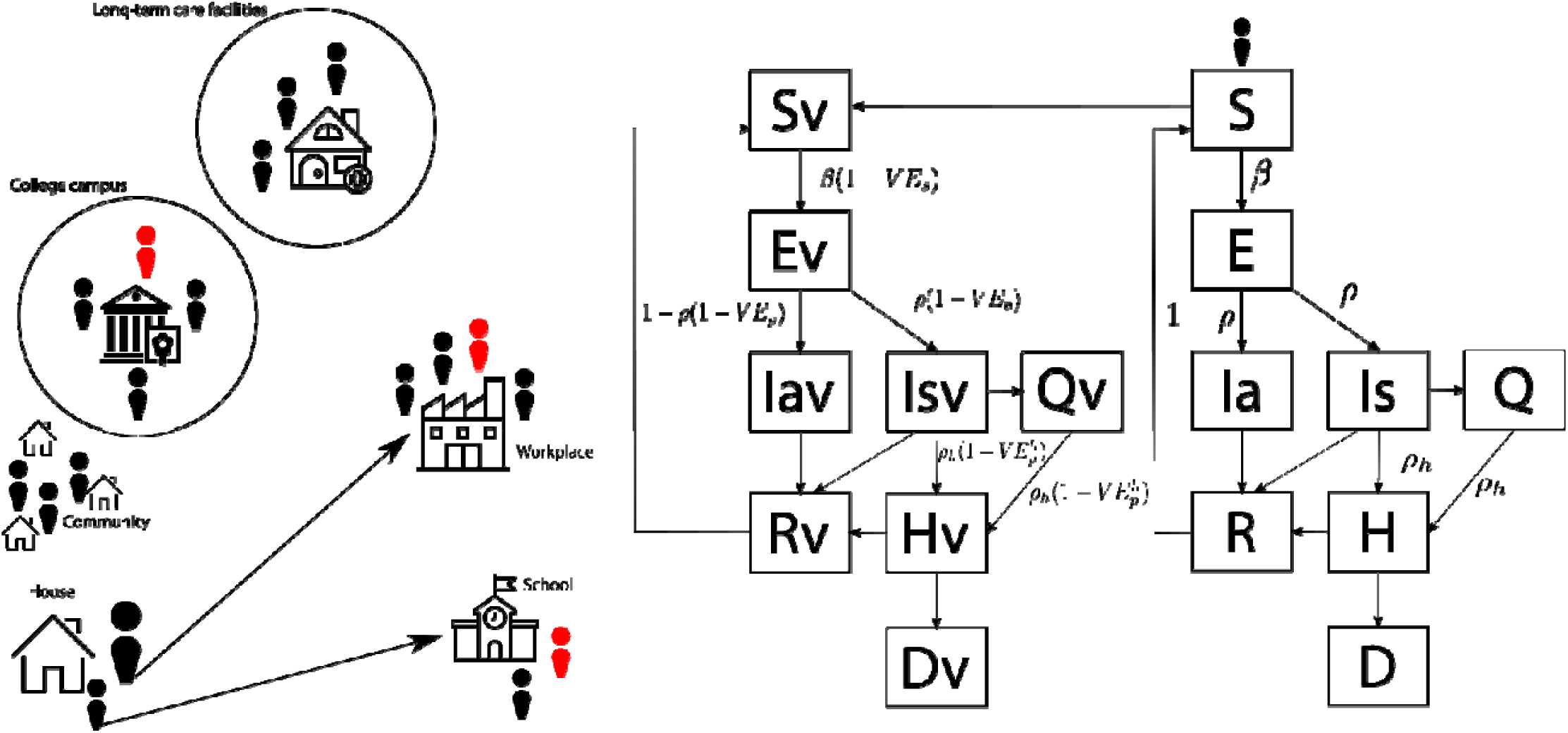
Model framework and individual transitions for vaccinated and unvaccinated agents.

We calibrated the model in a two-step fashion. First, we fitted parameters related to transmission of the original lineage of the virus by contrasting the model outputs to daily incidence of deaths reported from March 2020 to July 2021. Then, drawing from this calibrated distribution of parameters, we fitted parameters related to the transmissibility, immune escape (to immunity from natural infection), and introduction of the alpha, gamma, and B.1621 variants. In this step, we calibrated the model parameters to the daily incidence of deaths and the preliminary dominance data obtained from samples for epidemiologic weeks 12 to 22. Finally, the model was validated against the only available population-based seroprevalence study (conducted on November 2020 [21]) and daily ICU requests provided by the city’s Secretary of Health. Parameters of the delta variant were assumed as the median value of estimates from other studies [28].

### Variant’s parameters and importation

To calibrate the parameters related to transmission and immunity escape for the different variants, we used the ranges reported in the literature (Table S2). The date of introduction of variants was estimated using reported cases from international travelers and the prevalence of variants in their countries of origin [29], as well as the presence of the emerging B.1.621 variant in the country of origin [6]. Hence, the total number of imported infections from variant ‘*v*’ can be estimated as

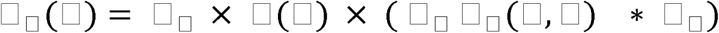

where *N(t)* is the total imports detected in the city for time *‘t’*;□_□_ (□,□) is the dominance of variant *‘v’* in country *‘c’* at time ‘*t*’ as reported at [22] as of June 30th, 2021; □_□_ is the overall proportion of importations detected in the city from country ‘*c*’; and □_□_ is a scaling factor estimated using daily deaths and dominance data from sequenced samples.

### Vaccination

The model uses an “*all-or-nothing”* assumption regarding the effect of vaccination, which means that the vaccine provides no protection at all to a fraction of the vaccinated persons and perfect lifetime immunity to the rest of the vaccinated ones [30]. Other studies have reported contrasting this modelling approach with one using a *“leaky”* assumption about the effect of vaccination, and have not found substantial differences for their projections about the potential impact of different vaccination strategies against SARS-CoV-2 [31].

### Vaccination parameters

based on the vaccination efficacy (VE) against disease (□□_□□_), we used a function of both the efficacy against infection □□_□_ and against the progression from infection to disease □□_□_, which under a multiplicative and independent relationship can be expressed as □□_□□_ = *1* − (*1* − □□_□_)(*1* − □□_□_) [32]. The efficacy against hospitalization could also be expressed as □□^□^_□□_ = *1* − (*1* − □□_□_) (*1* − □□_□_) (*1* − □□^□^_□_). We used reported □□_□□_ and □□^□^_□□_ to calculate □□_□_ and □□^□^_□_. Given the uncertainty about the vaccine efficacy against infection, we assumed different values of □□_□_ (equal to 50% or 100% of □□_□□_).

We included four vaccines currently used in Colombia, namely, Oxford/AstraZeneca (AZD1222), Pfizer-BioNTech BNT162b2, CoronaVac/Sinovac, and Janssen (J&J) Ad26.COV2.S. We used vaccine efficacy as reported from the clinical trials, except when the trial had not enough power to report efficacy (i.e. severe disease, death) or estimates were not available, in which case, results from observational studies were used. The point estimate and confidence intervals for the protection against symptomatic, and severe disease are listed in Table S1.

We considered that the protective effect of vaccines starts after a period that can vary between 7 and 15 days after the first and the second dose according to what is reported for each vaccine (Refs in Table S1). We assumed that vaccines have some level of protection against infection. Given the uncertainty in this regard, in our main analyses we assumed that protection against infection is 50% of that reported for symptomatic disease. We also considered an alternative scenario of efficacy against infection being 100% of the efficacy against symptomatic disease. The arrival of each type of vaccine and the maximum number of vaccines delivered each day were determined by the vaccination delivery reports by the country’s Ministry of Health [25], as shown in Fig S1. To project the future delivery of vaccines, we assumed constant availability with the proportion of vaccine types determined by the current one. In terms of the daily capacity of vaccine administration, we assumed that the current daily vaccination rates would remain until the target population is completely vaccinated. We also assume a probability of vaccine uptake of 90%.

Of note, we have assumed that VE against hospitalization for the alpha variant with one dose is substantially different compared to the same VE for the delta variant. For this parameter we have used VE as vaccine effectiveness reported, which has not been peer-reviewed [33].

### Simulation scenarios

We assessed the potential impact of delta variant introduction in the city by calibrating the transmission model to current patterns and projecting the number of deaths and the demand for ICU beds, from August to November 2021. The baseline scenario used the calibrated parameters for alpha, gamma, and B.1.621. For delta, we assumed the median value of studies reported in the literature [28]. The scaling factor of imports of delta was assumed to be a middle point similar to values calibrated to the alpha and gamma variants.

We simulated a set of scenarios to calculate the impact of mobility and the timing of the introduction of the delta variant. To evaluate the potential impact of mobility and contacts on the future dynamics of COVID-19 due to the delta variant, we simulated two different scenarios of mobility and contacts. The baseline scenario assumed the current levels of mobility and moderate contacts. We also considered a scenario of high mobility and high contacts, which assumed that contacts increased to the levels observed during December 2020. We evaluated two additional scenarios for the timing of the introduction of the delta variant: 30 or 60 days later than our baseline estimates.

We simulated alternative scenarios of vaccination considering: the increase in the vaccination capacity from 50,000 to 100,000 vaccines per day, a delivery with no age-based prioritization for future administration in contrast with the current age-prioritized strategy, and the postponing of the second dose of the Pfizer BioNTech’s BNT162b2 mRNA vaccine (from 21 to 84 days). We calculated the effect of these administration strategies on the cumulative number of deaths and ICU beds required from August 1st to November 1st, 2021, and the differences with the baseline scenario.

## Supplementary material

**Table S1.**
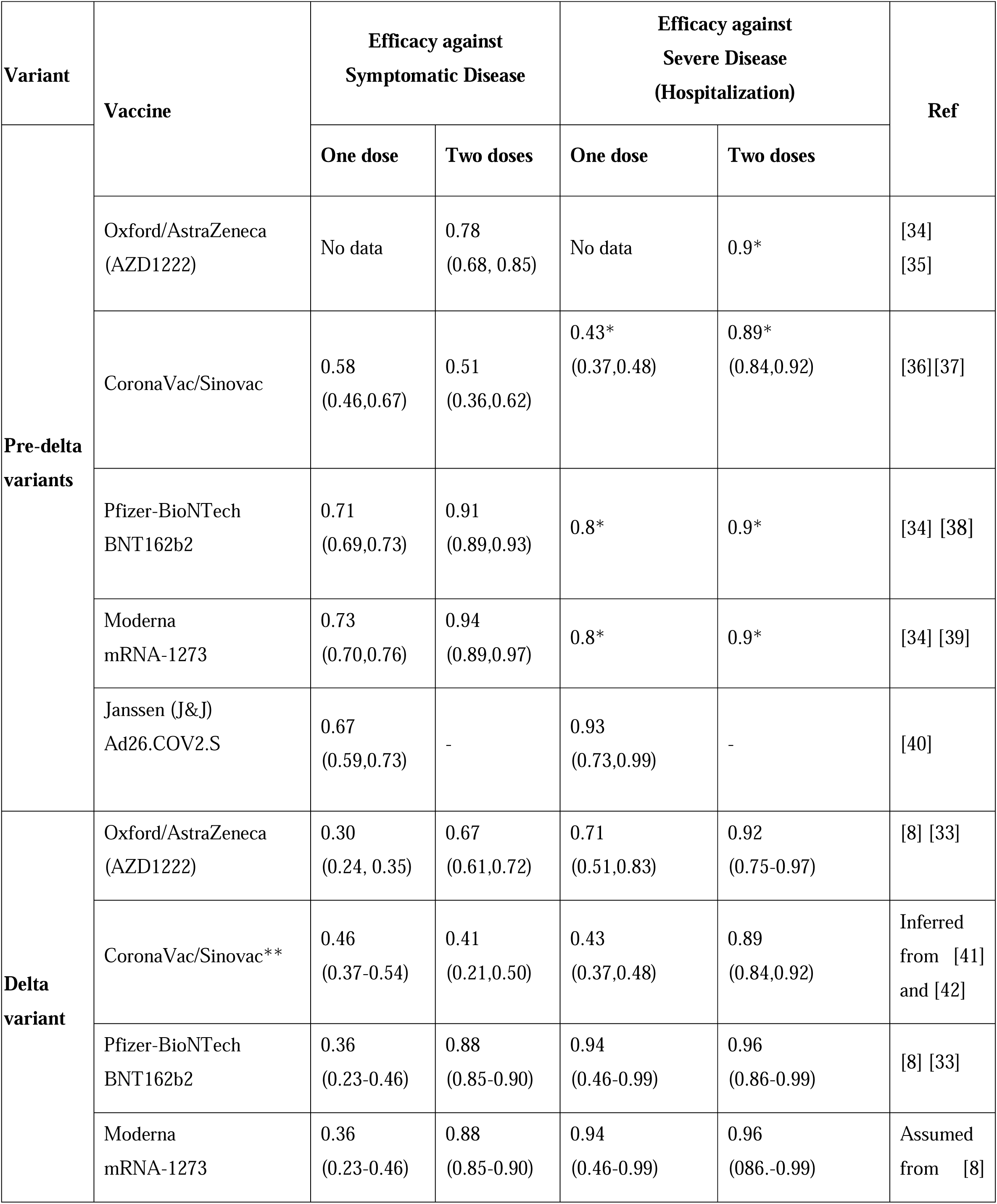
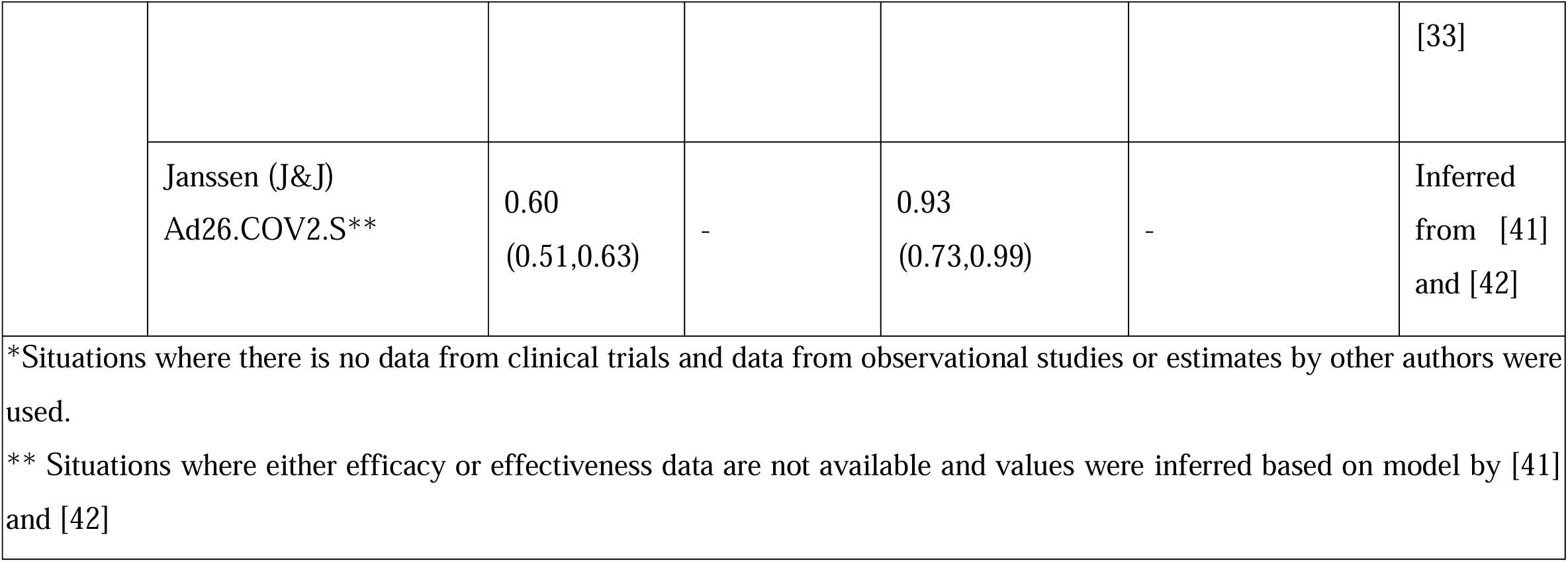
Parameter values for vaccine efficacy.

**Table S2.** Parameter bounds for characteristics of variants included in the calibration.

| **Variant** | **Transmission** | **Immune Escape** | **Date first reported in Colombia** | **Source** |
| --- | --- | --- | --- | --- |
| Alpha (B.1.1.7) | 1.5 | 0.00 - 0.05 | 2021-03-10 | [43] |
| Gamma (P.1) | 1.7 - 2.4 | 0.21 - 0.46 | 2021-03-24 | [44] |
| Delta (B.1.617.2) | 2.10 - 2.46 | 0.10 - 0.55 | 2021-07-24 | [28] |
| B.1621 | 1.0 - 2.0 | 0.0 - 0.5 | 2021-03-14 | Used as range for estimation within the model |

**Table S3.**
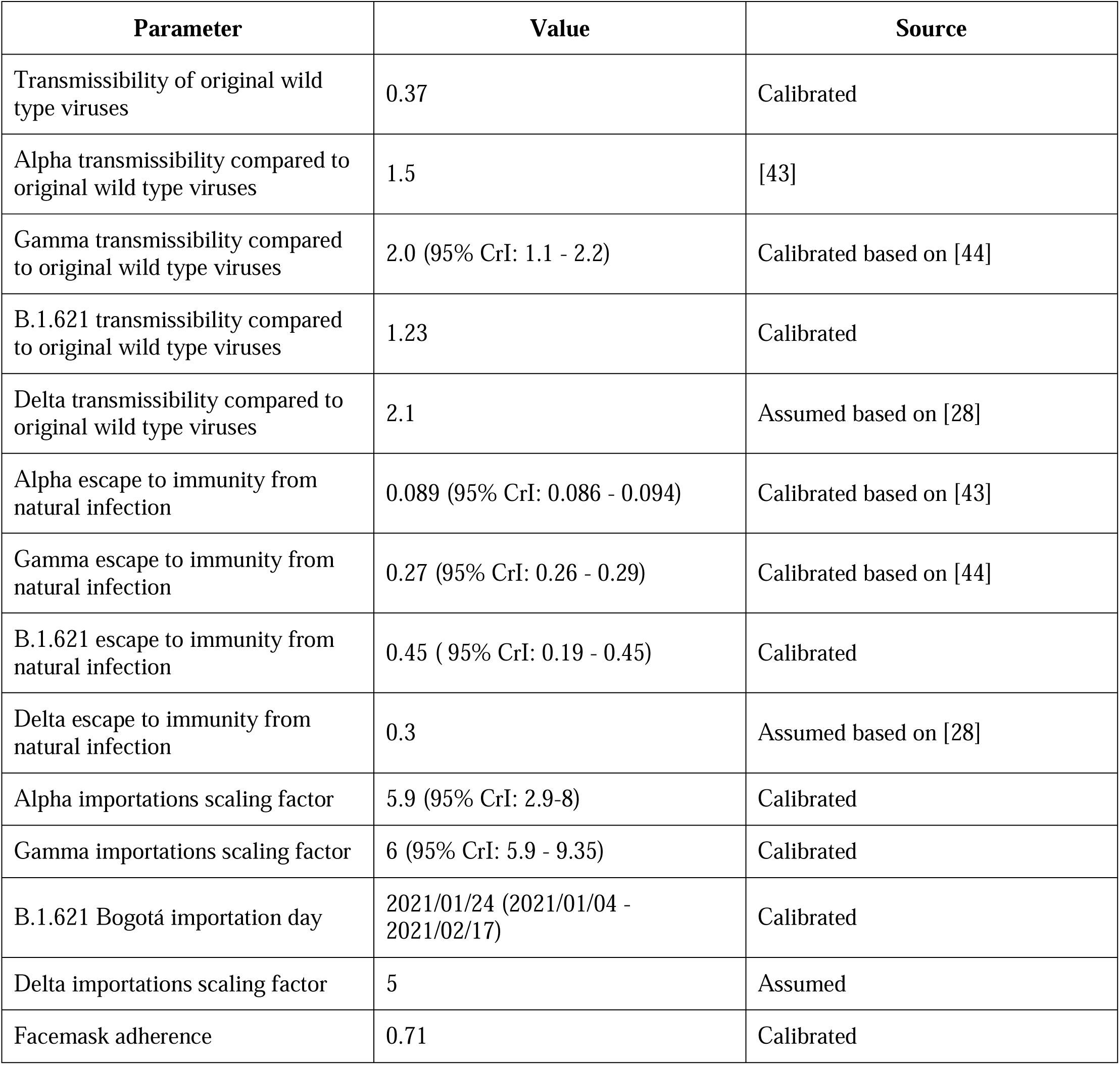
Calibrated parameters.

| Parameter | Value | Source |
| --- | --- | --- |
| Transmissibility of original wild type viruses | 0.37 | Calibrated |
| Alpha transmissibility compared to original wild type viruses | 1.5 | [43] |
| Gamma transmissibility compared to original wild type viruses | 2.0 (95% CrI: 1.1 - 2.2) | Calibrated based on [44] |
| B.1.621 transmissibility compared to original wild type viruses | 1.23 | Calibrated |
| Delta transmissibility compared to original wild type viruses | 2.1 | Assumed based on [28] |
| Alpha escape to immunity from natural infection | 0.089 (95% CrI: 0.086 - 0.094) | Calibrated based on [43] |
| Gamma escape to immunity from natural infection | 0.27 (95% CrI: 0.26 - 0.29) | Calibrated based on [44] |
| B.1.621 escape to immunity from natural infection | 0.45 ( 95% CrI: 0.19 - 0.45) | Calibrated |
| Delta escape to immunity from natural infection | 0.3 | Assumed based on [28] |
| Alpha importations scaling factor | 5.9 (95% CrI: 2.9-8) | Calibrated |
| Gamma importations scaling factor | 6 (95% CrI: 5.9 - 9.35) | Calibrated |
| B.1.621 Bogotá importation day | 2021/01/24 (2021/01/04 - 2021/02/17) | Calibrated |
| Delta importations scaling factor | 5 | Assumed |
| Facemask adherence | 0.71 | Calibrated |

**Fig S1.**
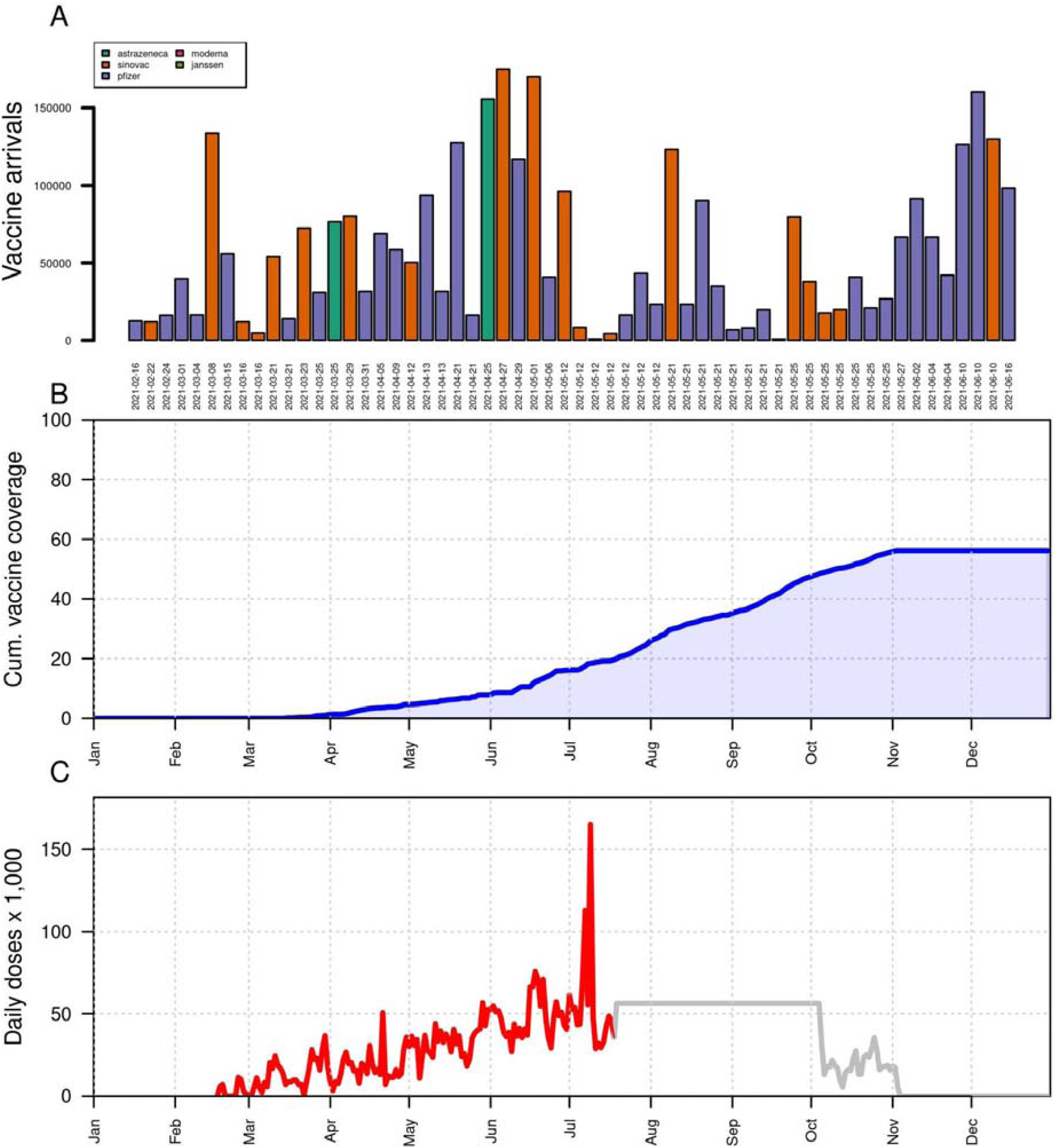
Vaccine stock and coverage in Bogotá. Panel A shows the arrival of vaccines by date and type of vaccine, panel B shows the projected coverage of full vaccination (fully vaccinated individuals divided by the total population of the city), and panel C shows daily vaccines administered in the model (gray) and data (red).

**Fig S2.**
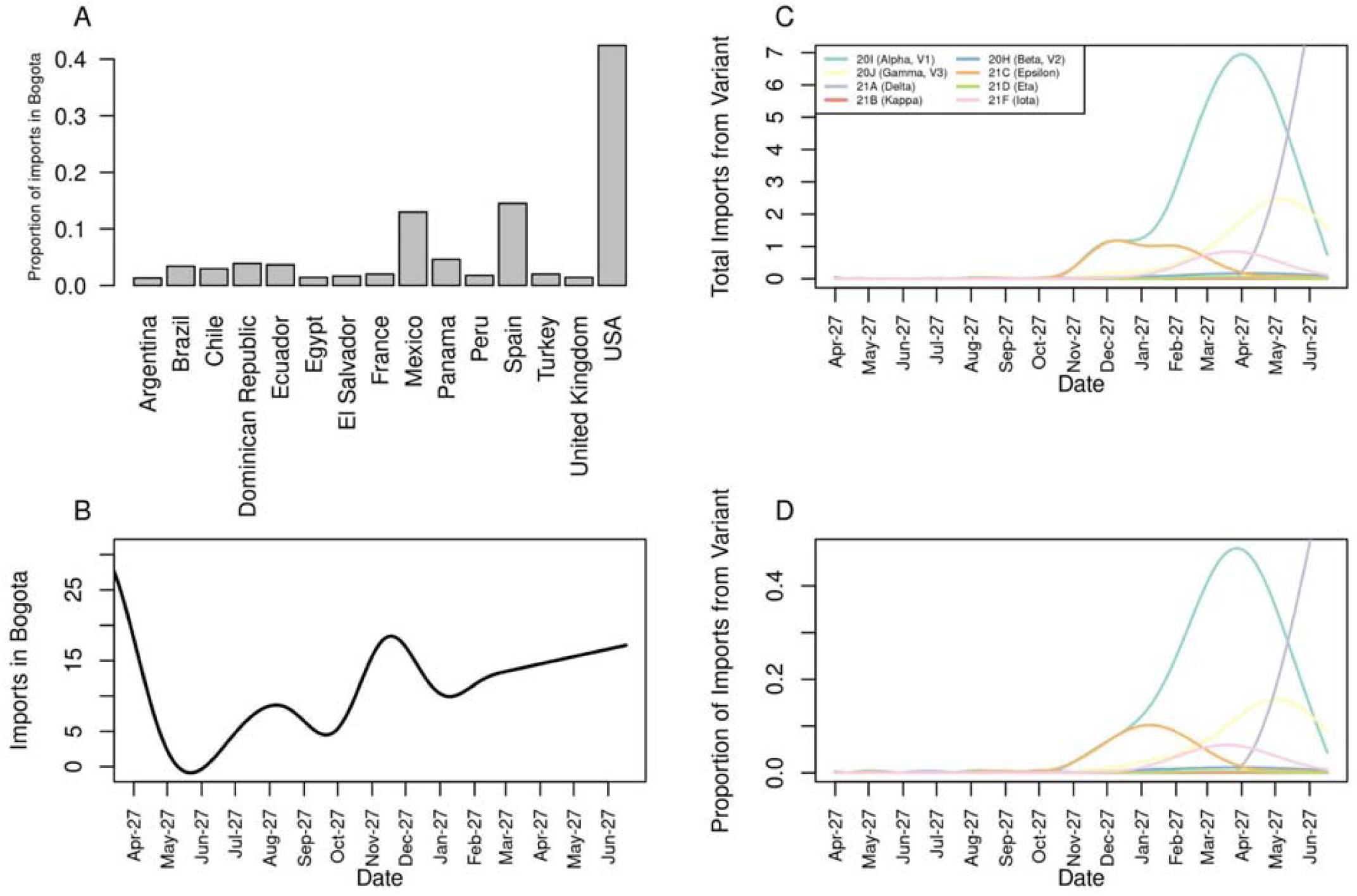
Importation of SARS-CoV-2 infections to Bogotá by variants and country of origin. Panel A shows the proportion of countries of origin from detected importations in the city of Bogotá, panel B shows a smoothed line of the trend of imports in the city, panel C shows the baseline estimated imports for each variant in the city, and panel D shows the proportion of the imports from each variant, according to prevalence in each country of origin.

**Fig S3.**
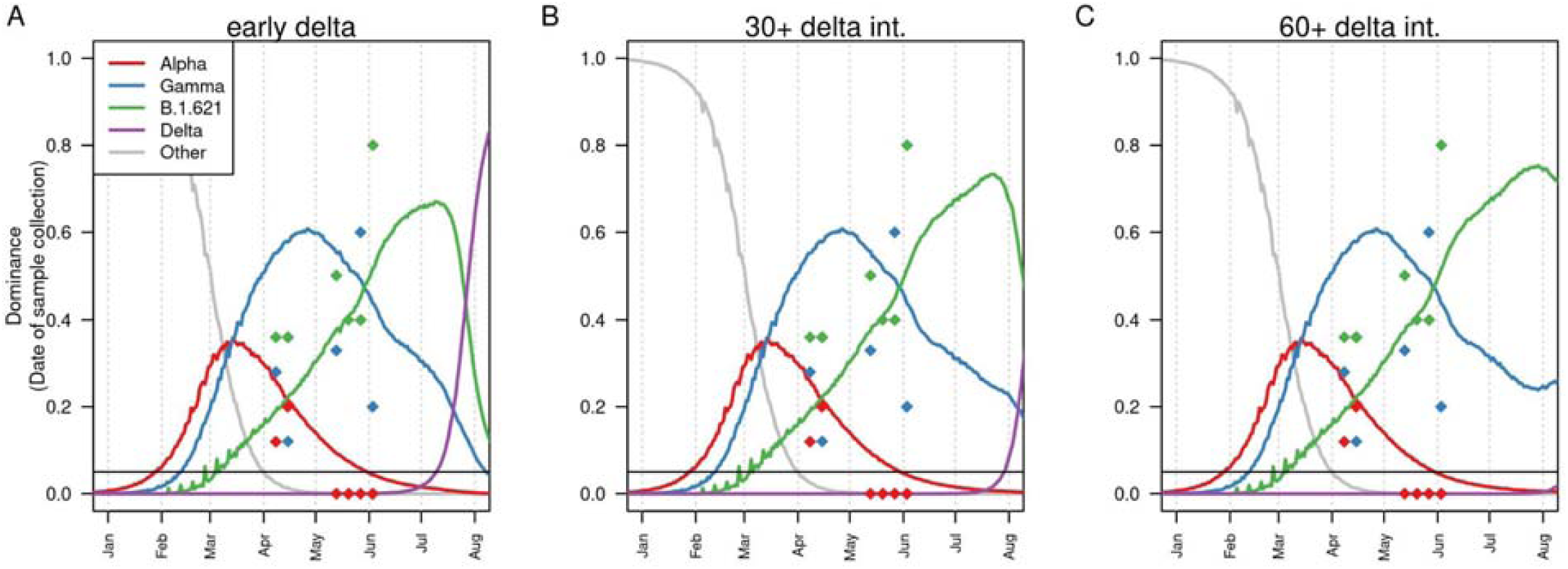
Model fit to dominance of variants. Panel A shows the dominance of different variants in the city for the scenario of early introduction of delta, panel B shows the dominance of variants for the scenario of delta 30+ (30 days delayed), and panel C shows the dominance of variants for the scenario of delta 60+(60 days delayed). Each line shows the dominance of each variant as estimated by the model. Diamonds show the data on dominance from these variants. Date of sample collection calculated as 7 days after infection.

**Fig S4.**
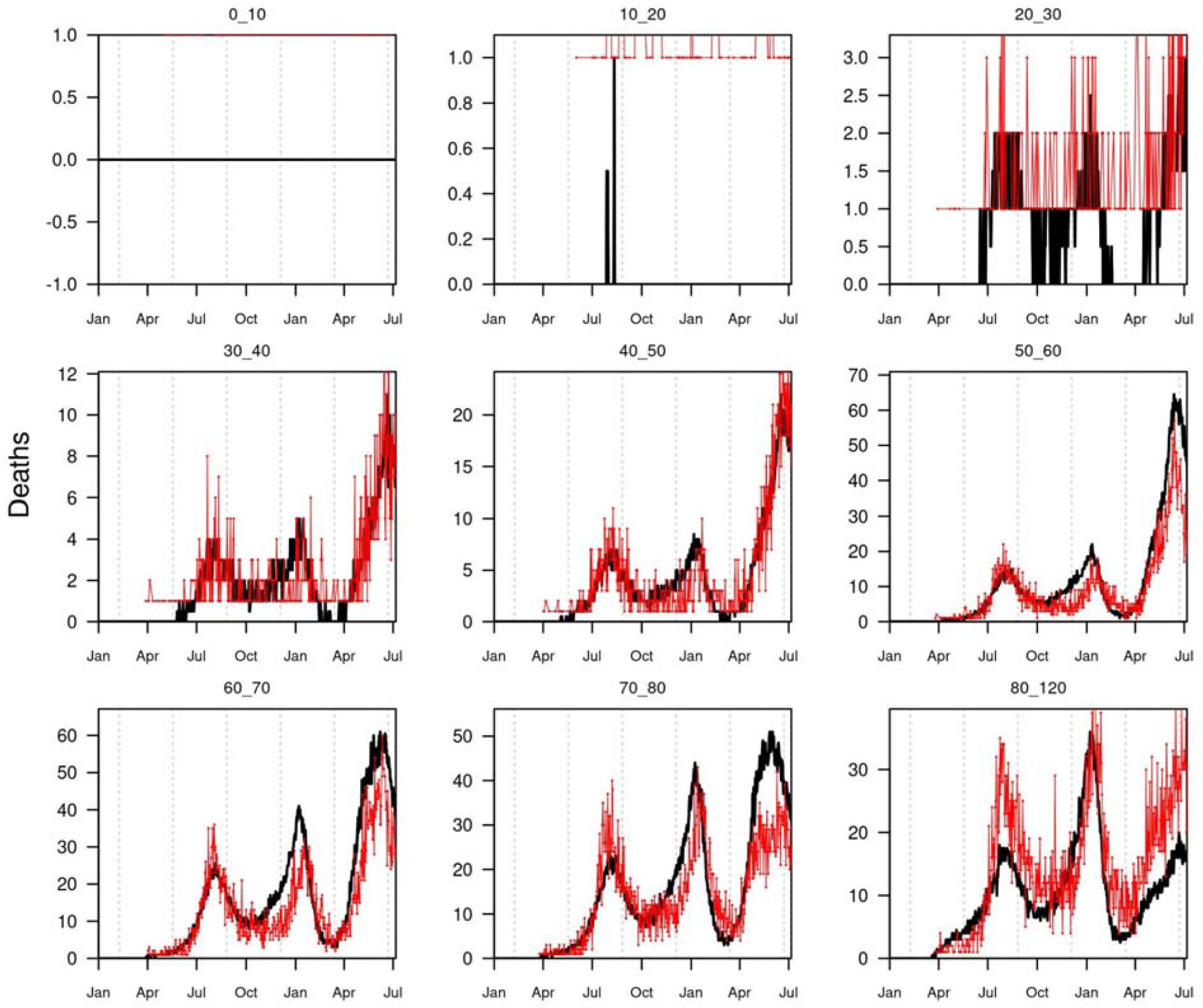
Model fit to age-specific deaths. Black lines show the model daily number of deaths, and red dots and lines show the reported daily deaths for each age group.

**Fig S5.**
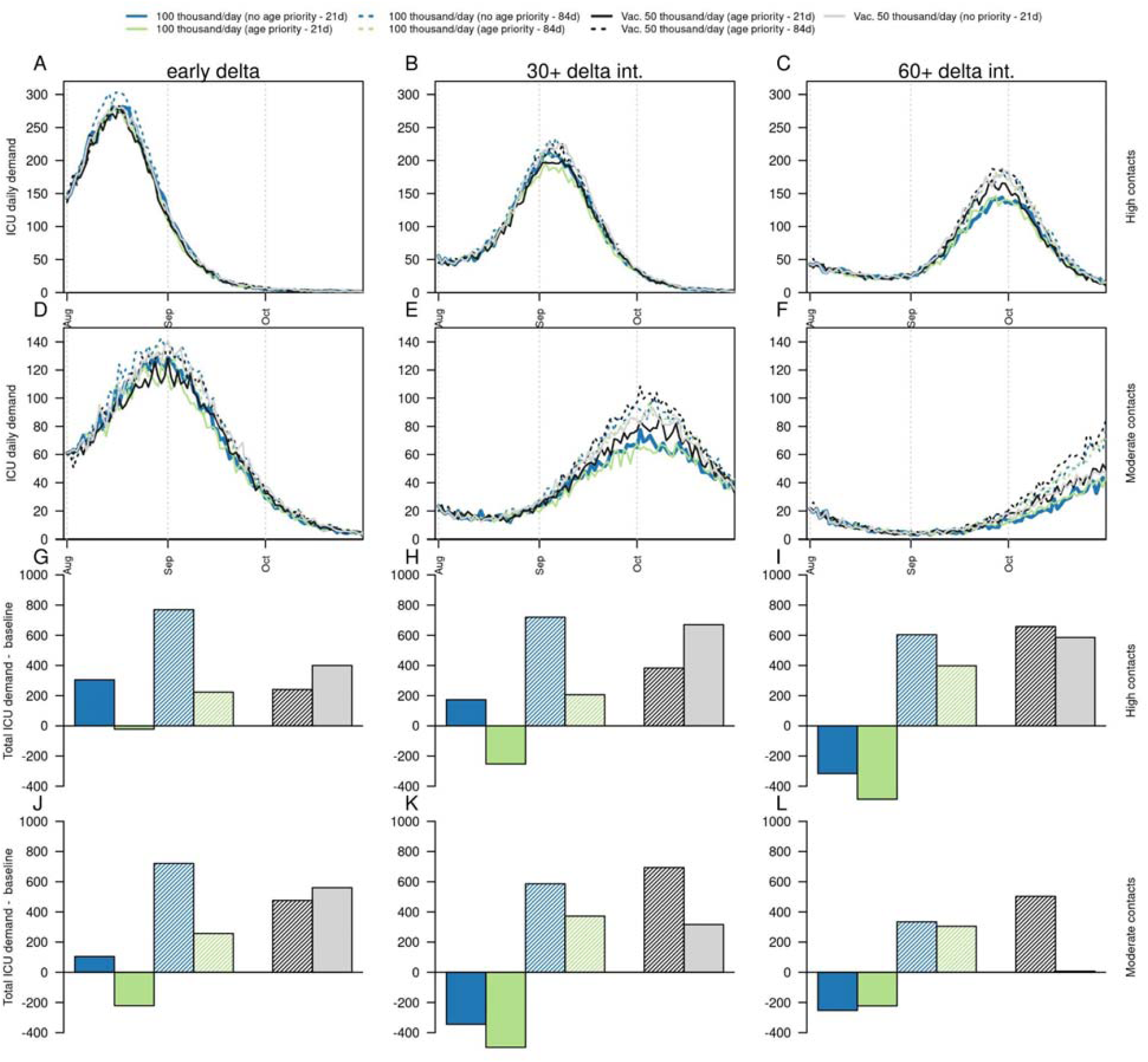
Projections of potential impact on ICU beds demand of delta variant on a fourth SARS-CoV-2 wave in Bogotá, according to timing of delta introduction, level of social mixing, and vaccination strategies. Columns show the timing of delta introduction, defined as early (calibrated), 30+ delta (delayed 30 days), and 60+ delta (delayed 60 days). Panels A-C show the daily number of ICU beds demanded under a scenario of high social mixing, panels D-F show the daily number of ICU beds demanded under a scenario of moderate social mixing, panels G-I show the difference in the cumulative number of ICU beds demanded between alternative vaccination strategies and the baseline scenario (50 thousand vaccines/day with age prioritization and non-postponed second dose of the Pfizer vaccine) with high social mixing, and panels J-L show the difference in the cumulative number of ICU beds between alternative vaccination strategies and the baseline scenario (50 thousand vaccines/day with age prioritization and non-postponed second dose) with moderate social mixing. Black line shows the baseline scenario of mobility and current vaccination strategy. Dashed lines show the impact of increasing the interval between doses to 84 days for the Pfizer vaccine. Blue colors show the impact of increased vaccination rates (100,000/day) without age priority. Green colors show the impact of increased vaccination rates (100,000/day) with age priority. Gray colors show the impact of baseline vaccination rates without age priority.

**Fig S6.**
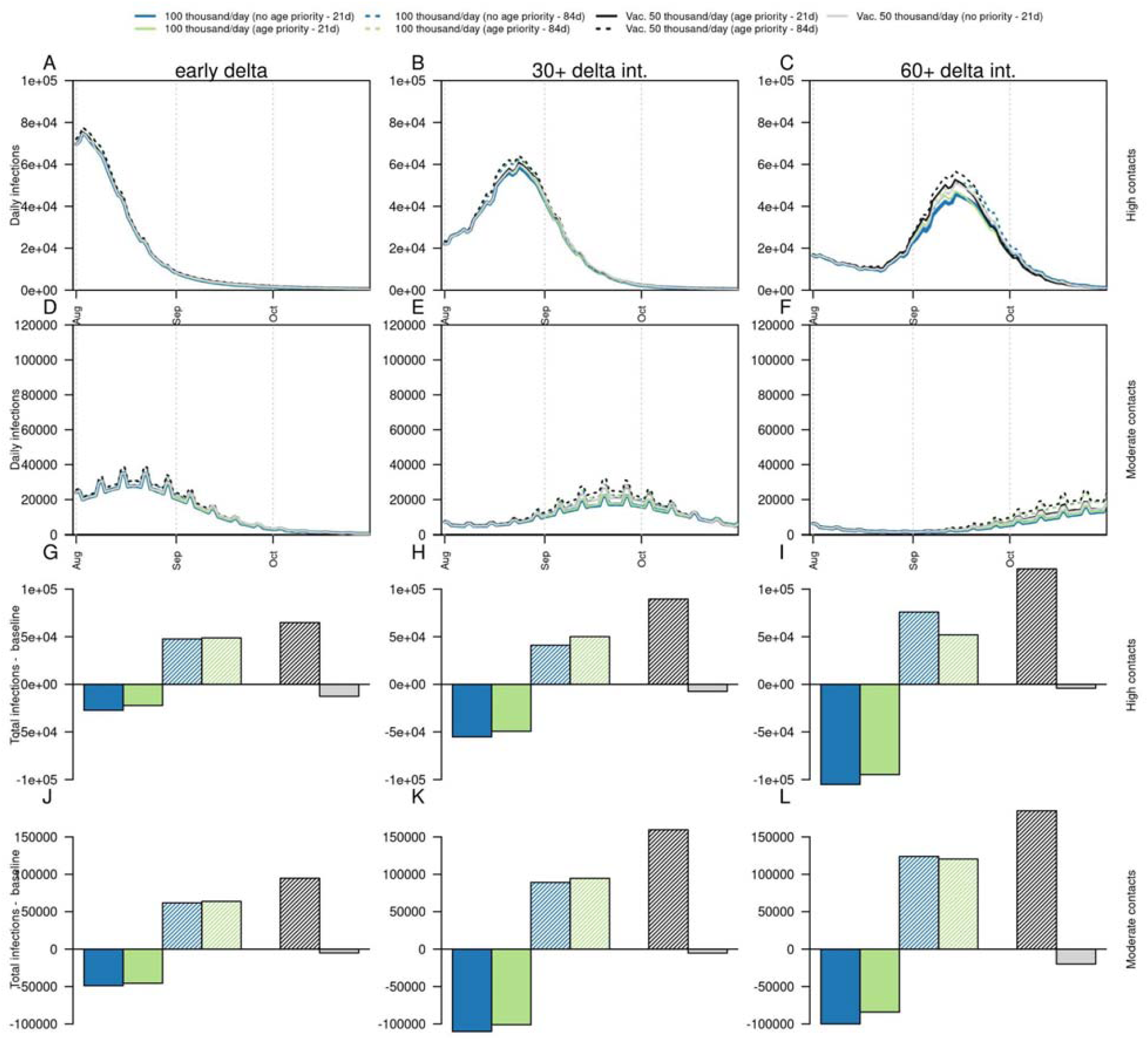
Projections of potential impact on infections of delta variant on a fourth SARS-CoV-2 wave in Bogotá, according to timing of delta introduction, level of social mixing, and vaccination strategies, under an alternative scenario of VE against infection (50% of VE against symptomatic disease). Columns show the timing of delta introduction, defined as early (calibrated), 30+ delta (delayed 30 days), and 60+ delta (delayed 60 days). Panels A-C show the daily number of infections under a scenario of high social mixing, panels D-F show the daily number of infections under a scenario of moderate social mixing, panels G-I show the difference in the cumulative number of infections between alternative vaccination strategies and the baseline scenario (50 thousand vaccines/day with age prioritization and non-postponed second dose of the Pfizer vaccine) with high social mixing, and panels J-L show the difference in the cumulative number of infections between alternative vaccination strategies and the baseline scenario (50 thousand vaccines/day with age prioritization and non-postponed second dose of the Pfizer vaccine) with moderate social mixing. Black line shows the baseline scenario of mobility and current vaccination strategy. Dashed lines show the impact of increasing the interval between doses to 84 days for the Pfizer vaccine. Blue colors show the impact of increased vaccination rates (100,000/day) without age priority. Green colors show the impact of increased vaccination rates (100,000/day) with age priority. Gray colors show the impact of baseline vaccination rates without age priority.

**Fig S7.**
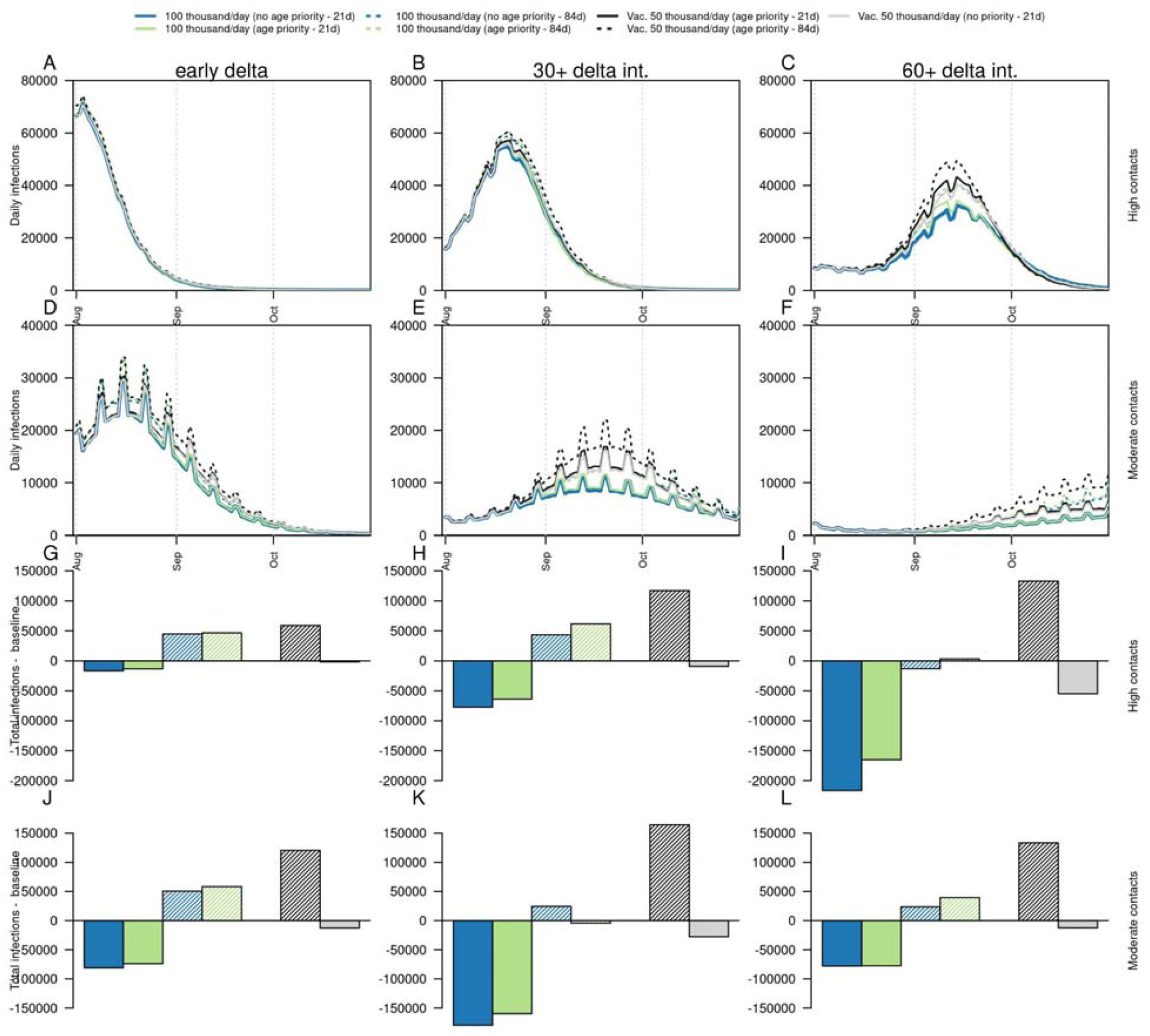
Projections of potential impact on infections of delta variant on a fourth SARS-CoV-2 wave in Bogotá, according to timing of delta introduction, level of social mixing, and vaccination strategies, under an alternative scenario of VE against infection (100% of VE against symptoms). Columns show the timing of delta introduction, defined as early (calibrated), 30+ delta (delayed 30 days), and 60+ delta (delayed 60 days). Panels A-C show the number of infections under a scenario of high social mixing, panels D-F show the number of infections under a scenario of moderate social mixing, panels G-I show the difference in the cumulative number of infections between alternative vaccination strategies and the baseline scenario (50 thousand vaccines/day with age prioritization and non-postponed second dose of the Pfizer vaccine) with high social mixing, and panels J-L show the difference in the cumulative number of infections between alternative vaccination strategies and the baseline scenario (50 thousand vaccines/day with age prioritization and non-postponed second dose) with moderate social mixing. Black line shows the baseline scenario of mobility and current vaccination strategy. Dashed lines show the impact of increasing the interval between doses to 84 days for the Pfizer vaccine. Blue colors show the impact of increased vaccination rates (100,000/day) without age priority. Green colors show the impact of increased vaccination rates (100,000/day) with age priority. Gray colors show the impact of baseline vaccination rates without age priority.

**Fig S8.**
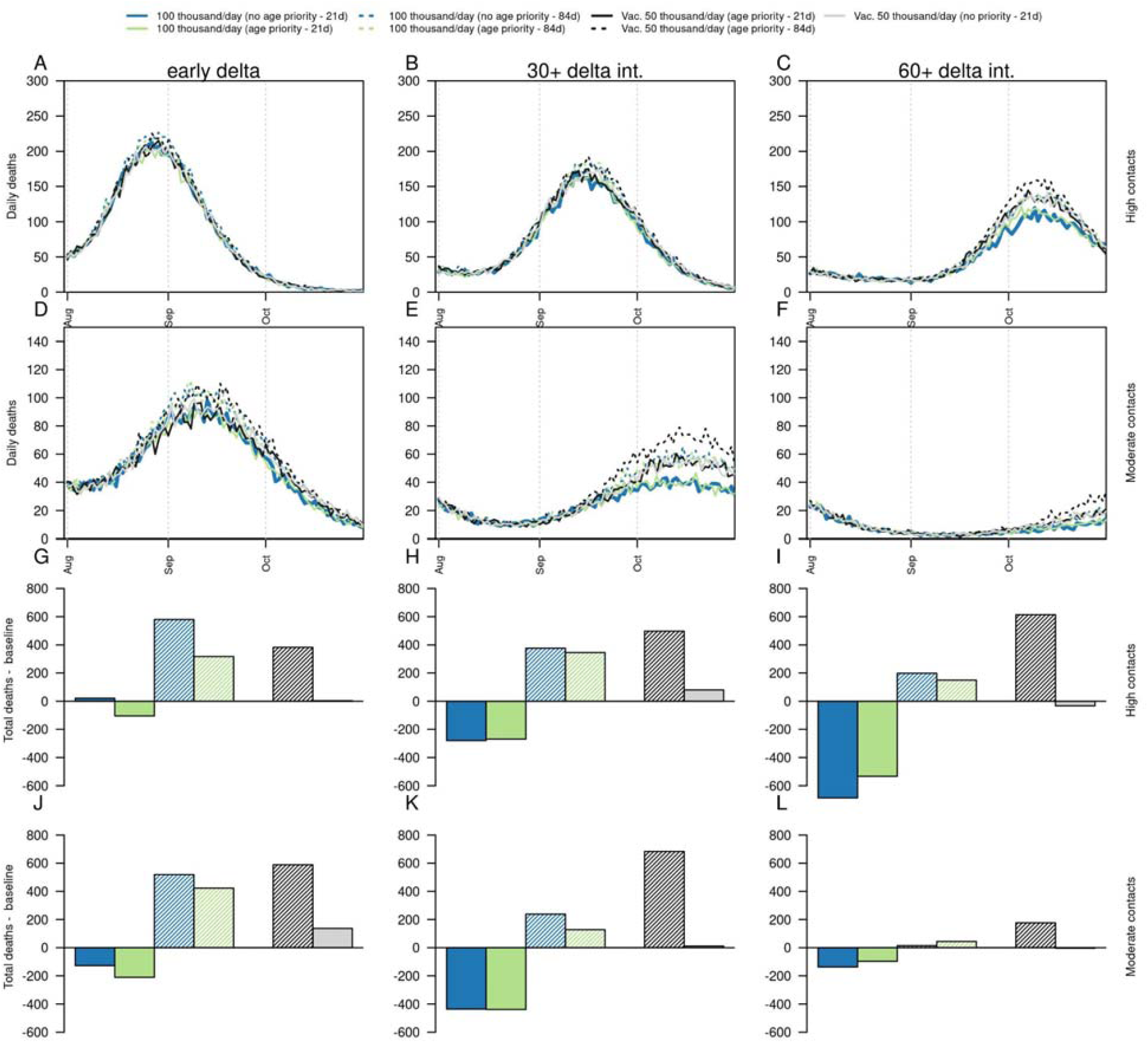
Projections of potential impact on deaths of delta variant on a fourth SARS-CoV-2 wave in Bogotá, according to timing of delta introduction, level of social mixing, and vaccination strategies, under an alternative scenario of VE against infection (100% of VE against symptomatic disease). Columns show the timing of delta introduction, defined as early (calibrated), 30+ delta (delayed 30 days), and 60+ delta (delayed 60 days). Panels A-C show the daily number of deaths under a scenario of high social mixing, panels D-F show the daily number of deaths under a scenario of moderate social mixing, panels G-I show the difference in the cumulative number of deaths between alternative vaccination strategies and the baseline scenario (50 thousand vaccines/day with age prioritization and non-postponed second dose of the Pfizer vaccine) with high social mixing, and panels J-L show the difference in the cumulative number of deaths between alternative vaccination strategies and the baseline scenario (50 thousand vaccines/day with age prioritization and non-postponed second dose) with moderate social mixing. Black line shows the baseline scenario of mobility and current vaccination strategy. Dashed lines show the impact of increasing the interval between doses to 84 days for the Pfizer vaccine. Blue colors show the impact of increased vaccination rates (100,000/day) without age priority. Green colors show the impact of increased vaccination rates (100,000/day) with age priority. Gray colors show the impact of baseline vaccination rates without age priority.

**Fig S9.**
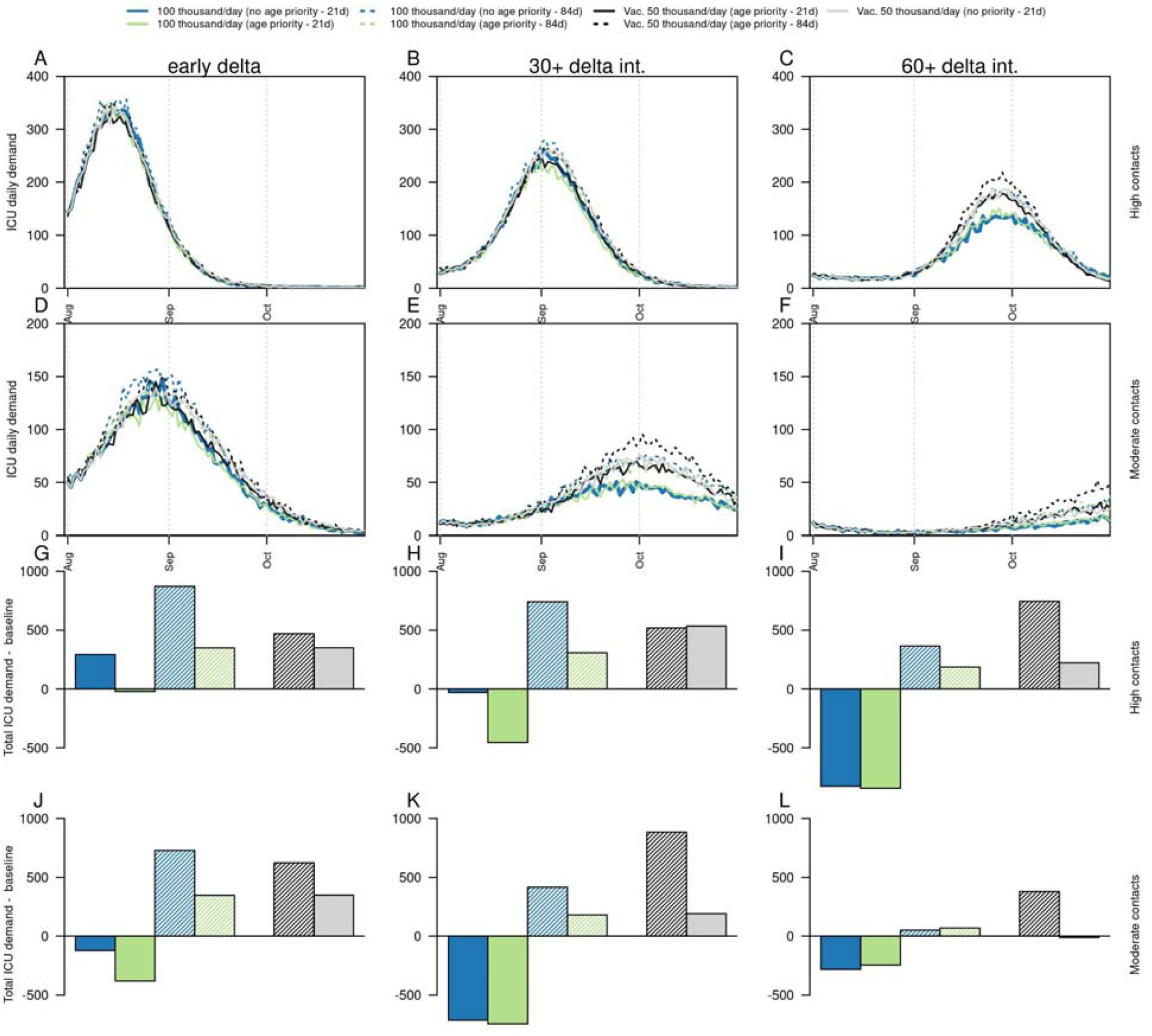
Projections of potential impact on ICU beds demand of delta variant on a fourth SARS-CoV-2 wave in Bogotá, according to timing of delta introduction, level of social mixing, and vaccination strategies, under an alternative scenario of VE against infection (100% of VE against symptoms). Columns show the timing of delta introduction, defined as early (calibrated), 30+ delta (delayed 30 days), and 60+ delta (delayed 60 days). Panels A-C show the daily number of ICU demand under a scenario of high social mixing, panels D-F show the daily number of ICU demand under a scenario of moderate social mixing, panels G-I show the difference in the cumulative number of ICU beds between alternative vaccination strategies and the baseline scenario (50 thousand vaccines/day with age prioritization and non-postponed second dose of the Pfizer vaccine) with high social mixing, and panels J-L show the difference in the cumulative number of ICU beds between alternative vaccination strategies and the baseline scenario (50 thousand vaccines/day with age prioritization and non-postponed second dose) with moderate social mixing. Black line shows the baseline scenario of mobility and current vaccination strategy. Dashed lines show the impact of increasing the interval between doses to 84 days for the Pfizer vaccine. Blue colors show the impact of increased vaccination rates (100,000/day) without age priority. Green colors show the impact of increased vaccination rates (100,000/day) with age priority. Gray colors show the impact of baseline vaccination rates without age priority.

